# Inflammation induced epigenetic activation of bivalent genes in osteoarthritic cartilage

**DOI:** 10.1101/2023.04.13.23288509

**Authors:** Hao Du, Yao Zhang, Xi Yu, Xuanhe You, Diwei Wu, Zhenyu Luo, Yongrui Cai, Hanpeng Lu, Zhixin Liao, Bi-Sen Ding, Ya Zhao, Yan Wang, Ke Xiao, Fan Yang, Fangji Gan, Ning Ning, Jiancheng Zeng, Zongke Zhou, Shishu Huang

**Author notes:** Corresponding author. (S.H.); (Z.Z.). These authors contributed equally to this work (Hao Du, Yao Zhang).

## Abstract

Osteoarthritis (OA) is the most prevalent joint disorder occurring with articular cartilage degradation, which includes a switch from an articular to a growth-plate chondrocyte phenotype. Epigenetics serves as a new therapeutic target but histone modification changes in OA remain elusive. Here, we investigated the profiles of four histone modifications in normal and OA chondrocytes. The repressive mark H3K27me3 was significantly lost in OA, associated with up-regulated gene expression. Surprisingly, many of these genes were occupied by both H3K27me3 and H3K4me3 in normal chondrocytes, showing a poised bivalent state. These bivalent genes are deemed to be activated during the hypertrophy of growth plate chondrocytes. Furthermore, inflammation induced the expression of demethylase KDM6B and decreased H3K27me3 level in OA chondrocytes, which was rescued by the KDM6B inhibitor GSK-J4. Altogether, our results suggest an inherited bivalent epigenetic signature on developmental genes that makes articular chondrocytes prone to hypertrophy and contribute to a promising epigenetic therapy for OA.

**The Paper Explained:** *Problem:* Osteoarthritis (OA) affects as much as 40% of the elderly population, representing the largest cause of age-related disability. The high susceptibility to OA suggests an intrinsic and systemic characteristic in articular chondrocytes that makes cartilage prone to degeneration.

*Results:* Epigenetic bivalent genes, which are occupied with both H3K27me3 and H3K4me3, are considered to poise expression of developmental genes. Surprisingly, we reported bivalency for hypertrophy related genes in normal articular chondrocytes. These bivalent genes need to be activated in growth plate chondrocytes for extracellular matrix degradation and ossification, but are left as a “bomb” for degeneration in articular chondrocytes. We further found that inflammation induced KDM6B remove H3K27me3 to activate hypertrophy related genes that promote OA.

*Impact:* Our results suggest an inherited epigenetic signature that makes articular chondrocytes prone to hypertrophy and ossification and contribute to a promising epigenetic therapy for OA.

## Introduction

Osteoarthritis (OA) is one of the most prevalent joint disorders that affects an estimated 300 million adults worldwide (Vos *et al*, 2017). OA is mainly characterized by chronic joint pain caused by the degradation of articular cartilage and inflammation of the synovium, which would induce age-related disability eventually. Current standard therapies for OA are limited, including pain amelioration and eventual total joint replacement (Bannuru *et al*, 2019), which do not prevent or restrain the loss of cartilage.

Up to date, many therapeutic strategies have focused on molecular pathways that are deregulated in OA. The relevant drugs include metalloproteinase and aggrecanase inhibitors to target catabolic enzymes responsible for degradation of articular cartilage, bisphosphonates and antiresorptive drugs to stabilize bone homeostasis, inducible nitric oxide synthase (iNOS) and NF-κB inhibitors to target inflammatory pathways (Karsdal *et al*, 2016). These drugs, however, have had limited preclinical and clinical success (Karsdal *et al*, 2016). One possible reason is that OA is a disease of complex pathology which involves multiple signaling pathways and these drugs target single molecular pathway only. In fact, most risk factors of OA, including aging, obesity, trauma and biomechanics, have been associated to epigenetic changes (Yang *et al*, 2021; Zhang *et al*, 2020; Ling & Rönn, 2019). Interestingly, epigenetic regulations can act on multiple gene programs at the same time, thus having the potential to reprogram the aberrant state of the cell. However, it is unknown whether epigenetic changes may play a common pathogenic role in the development of OA. As such, it is worth investigating epigenetic changes associated with OA, their potential impact on OA-associated gene expression, and disease pathology.

Epigenetics is defined as the changes ‘on top of’ (epi) the genome that influence the transcription of genes, which mainly imply DNA methylation and various histone modifications. Multiple studies have mapped global DNA methylation changes associated with OA (Taylor *et al*, 2015; Reynard, 2017) and provide an attractive therapeutic strategy (Smeriglio *et al*, 2020). Compared to DNA methylation, histone modifications are more diverse, complex and have close interactions with the former, providing broader therapeutic prospect (Grandi & Bhutani, 2020).

Histones are subject to post-translational modifications that include methylation, acetylation, phosphorylation and ubiquitylation, among others (Millán-Zambrano *et al*, 2022). Different histone modifications occupy distinct positions on the genome and confer distinct active or repressive transcription effect (Hyun *et al*, 2017). Histone 3 lysine 4 trimethylation (H3K4me3) marks active transcription and is highly enriched at the promoter region and transcription start site. H3K27me3 is a hallmark of transcriptional repression. H3K9me3 is a well-known indicator of silenced transcription and heterochromatin structure. Acetylation of H3K27 (H3K27ac) is the best-known epigenetic mark for active enhancers and also enriched in highly expressed genes. Promisingly, plenty of studies have revealed the involvement of histone modification enzymes in cartilage development and pathology (Dudakovic *et al*, 2015, 2; Camilleri *et al*, 2018, 2; Du *et al*, 2020, 2; Zhang *et al*, 2015, 3; Dai *et al*, 2017, 6; Monteagudo *et al*, 2017). While genome-wide changes of histone modifications associated with OA are still unknown.

In this study, we compared the genome-wide profiles of classic histone modifications in normal and OA chondrocytes and identified a significant decrease of repressive mark H3K27me3 in OA. This H3K27me3 loss derepressed gene expression, especially of bivalent genes, which are occupied by both H3K27me3 and H3K4me3. Interestingly, these genes are deemed to be activated during developmental chondrocyte hypertrophy. Further investigation revealed that inflammation induced histone demethylase KDM6B lead to H3K27me3 loss and can be an attractive therapeutic target for OA.

## Results

### The profiling of histone modifications in normal and OA chondrocytes

To gain a comprehensive view of the histone modification landscape in human articular chondrocytes and identify epigenetic changes associated with OA, we performed CUT&Tag and RNA-seq in normal and OA chondrocytes (Fig 1A). Two pairs of OA- Intact/Damaged samples were also included. OA-Intact cartilage showed slightly uneven surface compared to normal cartilage, whereas OA-Damaged cartilage exhibited severe erosion (Fig EV1A). We investigated the distribution of the activating promoter mark H3K4me3, the repressive mark H3K27me3, the promoter and enhancer mark H3K27ac, and the heterochromatin mark H3K9me3.

**Figure 1.**
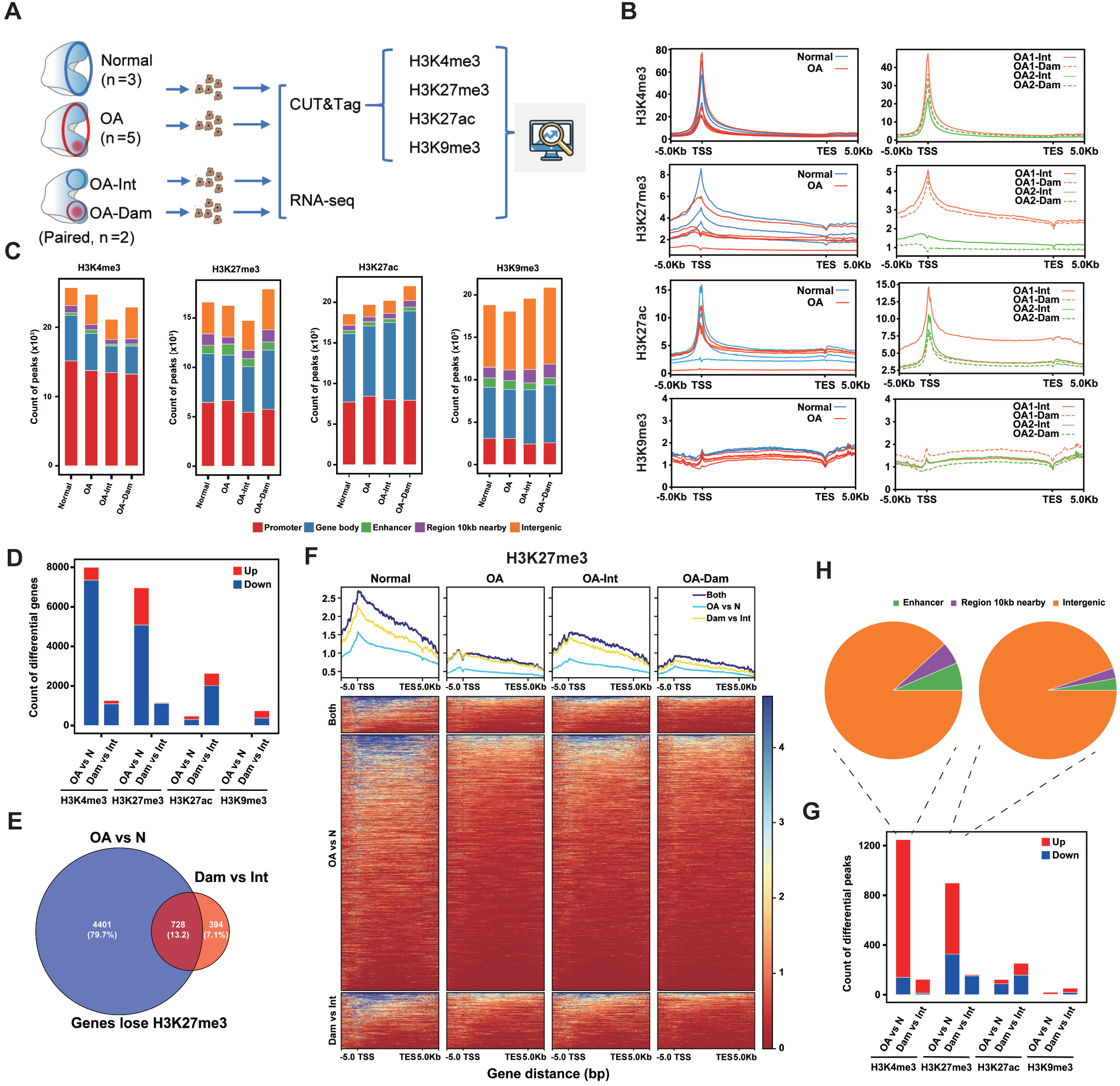
Histone modification profiling of normal and OA chondrocytes. (A) Schematic diagram of the experimental design. Chondrocytes from Normal (n=3), OA (n=5), OA-Intact and OA-Damaged (Paired, n=2) samples were used for CUT&Tag and RNA-seq. (B) Tag density of each histone mark across the gene bodies (+/-5kb) in Normal and OA (left), paired OA-intact and OA-damaged (right). (C) Genome-wide peak distribution of each modification in Normal, OA, OA-intact and OA- damaged samples. The numbers of peaks in promoter, gene body, enhancer, 10kb nearby gene body, and intergenic regions are shown. (D) The numbers of differential modified genes (log2FC>0.58, p-value<0.05) between Normal and OA, OA-intact and OA-damaged for each histone mark. (E) Venn diagram of genes which lose H3K27me3 in OA vs. Normal and OA-Damaged vs. OA-Intact. (F) Tag density and heatmap of H3K27me3 across the gene bodies (+/-5kb) of genes which loss H3K27me3 in OA vs. Normal, OA-Damaged vs. OA-Intact and both. (G) The numbers of differential peaks outside genes (log2FC>0.58, p-value<0.05) between Normal and OA, OA-intact and OA-damaged for each histone mark. (H) Distribution of up-regulated peaks outside genes for H3K4me3 and H3K27me3 in OA vs. Normal.

We first examined the tag density of each chromatin mark in genes. H3K4me3 and H3K27ac were highly enriched at the transcription start site (TSS) (Fig 1B and Fig EV1B). H3K27me3 were enriched at promoters and showed broader distribution than H3K4me3. Surprisingly, OA samples showed overall lower H3K27me3 compared to Normal (Fig 1B). The heterochromatin mark H3K9me3 showed low tag density on gene bodies in all samples. Subsequently, we used SEACR (Sparse Enrichment Analysis for CUT&RUN) script to identify genomic regions enriched for each chromatin mark.

About 20000 peaks were obtained for each group (Fig 1C) and 10000∼20000 peaks for each sample (Fig EV1C). The genome-wide peak distribution varied among different modifications but showed no significant difference in the four groups (Fig 1C). For H3K4me3, H3K27me3 and H3K27ac, most peaks were located in promoters and gene bodies. H3K9me3 peaks mainly located in intergenic regions and gene bodies (Fig 1C). The expected distribution of the four marks illustrated the reliance of our CUT&Tag experimental procedure.

As OA exhibited lower H3K27me3 in genes than Normal (Fig 1B), we wondered whether this change reflects an overall decrease or was restricted to a group of genes. To elucidate genes with differential H3K27me3 and other histone marks, we calculated peak strength that locate in each gene (TSS +/-2kb). Indeed, about 7000 genes were identified to have differential H3K27me3 strength in OA versus Normal, with more than 5000 genes lost H3K27me3 in OA (Fig 1D). Surprisingly, in Damaged versus Intact, over 1000 genes lost H3K27me3 and very few genes gain H3K27me3 (Fig 1D), in accordance with the overall decrease in tag density (Fig 1B). The dominance of H3K27me3 down-regulated genes in both compares indicated the importance of this H3K27me3 loss. Furthermore, the majority of genes that were found to lose H3K27me3 in Damaged versus Intact were also found in OA versus Normal (Fig 1E). These genes gradually lost H3K27me3 from Normal to OA-Intact to OA-Damaged (Fig 1F). For H3K4me3, more than 7000 genes showed a decrease in OA versus Normal, and over 1000 genes showed a decrease in Damaged versus Intact (Fig 1D and Fig EV2A). However, the decrease of H3K4me3 was weaker than H3K27me3 (Fig 1F and Fig EV2B). For H3K27ac and H3K9me3, relatively few differential genes were found in OA versus Normal (Fig 1D).

As the histone modification peaks, especially H3K9me3, also distributed in intergenic regions (Fig 1C), peaks outside genes with differential strength were calculated between OA and Normal, Damaged and Intact (Fig 1G). However, few differential H3K9me3 peaks were found in both compares. Surprisingly, most differential H3K4me3 peaks outside genes were up-regulated, in both OA versus Normal and Damaged versus Intact (Fig 1, G and H). Moreover, the up-regulated H3K4me3 peaks were enriched around centromere (Fig EV2C), while H3K27me3 didn’t show similar pattern (Fig EV2D), which needs further exploration.

Genome-wide correlation among all samples suggested that H3K27me3 was more effective than the other three marks to distinguish between OA and Normal (Fig EV3).

### Correlation between H3K27me3 loss and gene up-regulation in OA

To correlate histone modification to gene expression, transcriptomes of the corresponding samples were analyzed (Fig 2A). In OA versus Normal, 583 genes were down-regulated, and 297 genes were up-regulated. In Damaged versus Intact, 1327 genes were down-regulated, and 657 genes were up-regulated. First, we plotted the RNA expression levels against the modification levels in genes for normal samples (Fig 2B). This analysis indicated that gene expression positively correlated with H3K4me3 and H3K27ac in chondrocytes. The repressive mark H3K27me3 were associated with genes expressed at low levels and negatively correlated with the expression levels. The expected correlations further proved the reliability of our methods.

**Figure 2.**
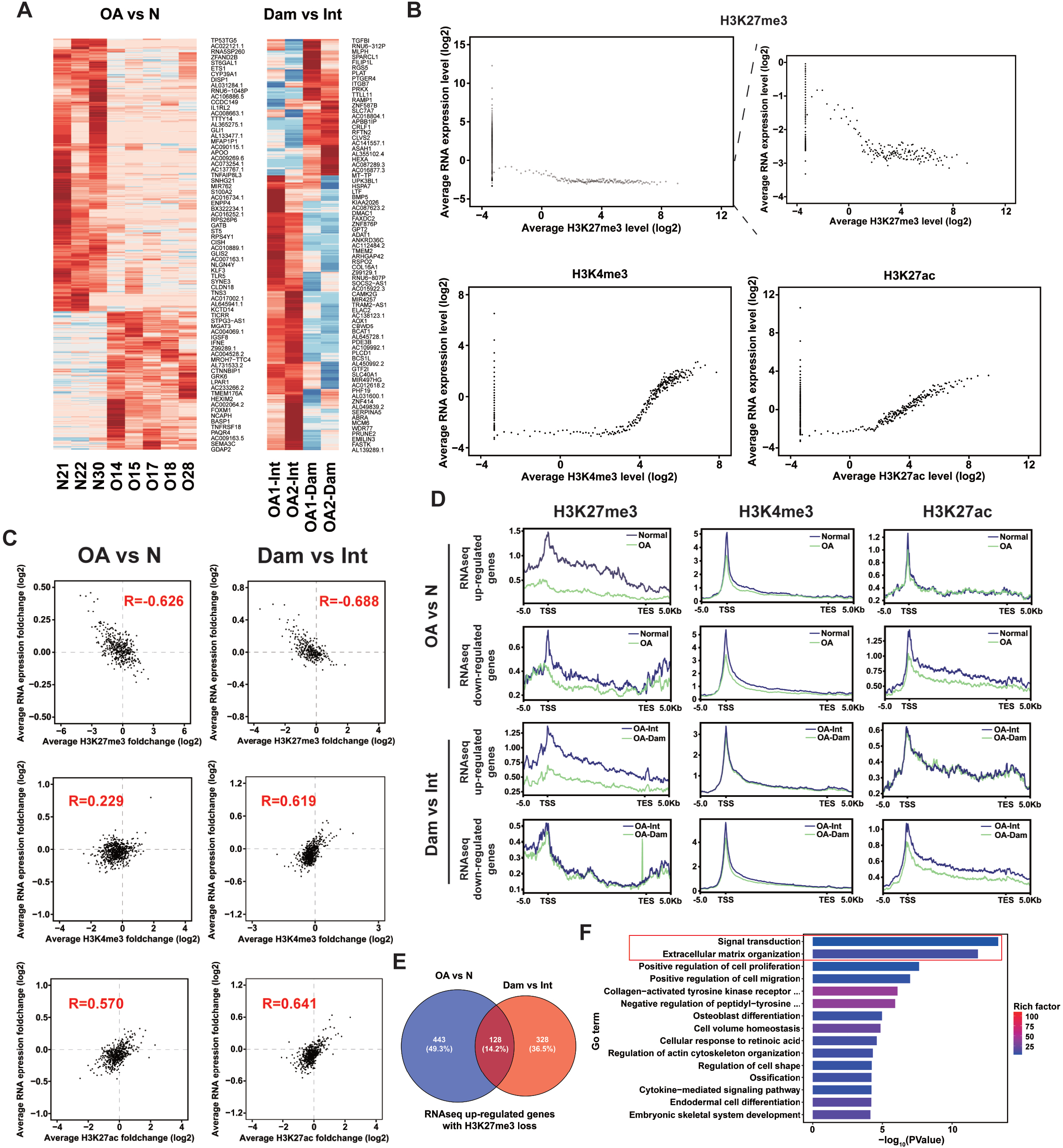
Correlation between histone modification and differential gene expression. (A) Heatmap of differential gene expression (log2FC>0.58) between Normal and OA, OA- intact and OA-damaged. (B) Correlation between each histone modification and gene expression in normal chondrocytes. Genes were grouped to 100 gene (one dot in the online supplemental figure) sets according to each histone modification level. (C) Pearson correlation between changes in histone modification and gene expression for OA vs. Normal and OA-Damaged vs. OA-Intact. The fold changes in both expression level and histone modification level were calculated for each gene. The genes were grouped into 100 gene sets according to their histone modification changes. (D) Tag density of H3K27me3, H3K4me3 and H3K27ac across the gene bodies (+/-5kb) of genes up-regulated or down-regulated expression in OA vs. Normal and OA-Damaged vs. OA-Intact. (E) Venn diagram of genes which lose H3K27me3 and gain expression in OA vs. Normal and OA-Damaged vs. OA-Intact. (F) GO terms with the highest enrichment of 128 genes which lose H3K27me3 and gain expression in both OA vs. Normal and OA-Damaged vs. OA-Intact.

To elucidate the correlation between modification and gene expression changes, the RNA expression fold changes were plotted against the histone modification fold changes in OA versus Normal and Damaged versus Intact (Fig 2C). The H3K27me3 modification changes inversely correlated with gene expression changes, and H3K27ac positively correlated with gene expression. H3K4me3 changes positively correlated with expression changes in Damaged versus Intact, but not in OA versus Normal. Overall, OA samples exhibited strongest correlation between decreased H3K27me3 and up- regulated gene expression (Fig 2C), consistent with the significant H3K27me3 loss in OA (Fig 1, D and F).

Additionally, we investigated the changes of histone modifications in up or down- regulated gene groups (Fig 2D). Surprisingly, H3K27me3 decreased markedly in up- regulated genes in both OA versus Normal and Damaged versus Intact, and H3K4me3 and H3K27ac level showed no significant change. This phenomenon indicated that the gene up-regulation in OA mainly depends on the removal of repressive mark H3K27me3 rather than the addition of activating marks. For the down-regulated genes, H3K27ac exhibited moderate decrease in both OA versus Normal and Damaged versus Intact (Fig 2D). Repressive mark H3K27me3 showed a slight decrease in OA versus Normal, which may result from the global H3K27me3 loss in OA (Fig 1, B and D). H3K4me3 showed a slight decrease in both down-regulated and up-regulated genes in OA versus Normal. This reflected an overall slight decrease of H3K4me3 (Fig 1D and Fig EV2B), which did not affect gene expression. Specifically, 443 genes were identified to lose H3K27me3 and gain expression in OA versus Normal, with 328 genes in Damaged versus Intact, and 128 genes in both compared groups (Fig 2E). The overlapped genes were highly enriched in GO terms like inflammatory signaling and extracellular matrix organization (Fig 2F), which are among the most typical changes in OA pathology, suggesting biological significance of H3K27me3 loss-mediated gene derepression in OA.

### H3K27me3 loss mediated up-regulation mainly occur on bivalent genes

As previously discovered, thousands of genes lost H3K27me3 in OA (Fig 1D) but only hundreds of them were up-regulated (Fig 2E). We wondered whether other histone marks contribute to this difference. Accordingly, we defined 12 chromatin states carrying distinct combinations of the four histone marks using ChromHMM (Ernst & Kellis, 2017) (Fig 3A). The enrichment of these chromatin states was examined at global level and in specific genomic features in three normal samples (Fig EV4A). Genome-wide correlation of chromatin states among all samples suggested better discrimination between Normal and OA samples than each single histone mark (Fig EV4B and S3). Then we defined the promoter state of each gene by the chromatin states of 2kb region around TSS (Fig 3B). The most prevalent promoter state was empty (state 12, 3) which include about 20000 genes in Normal. Other promoter states were activating (states 9, 10, 11), repressive (states 5, 6, 7) and bivalent (state 8). H3K27me3 exist in states 5, 6, 7 and 8. Surprisingly, genes with bivalent promoters (state 8) account for more than 2000 in Normal and decreased by half in OA (Fig 3B). As bivalent domains are well known in embryonic stem cells but not in terminal-differentiated cells, we wondered whether the observed bivalency came from mixed populations of cells that carry either repressive or activating marks at a given locus. However, the very low expression level of these genes in normal chondrocytes (Fig 3I) didn’t support the exist of activating cell population. Although terminally differentiated, normal articular chondrocytes still harbored plenty of bivalent genes.

**Figure 3.**
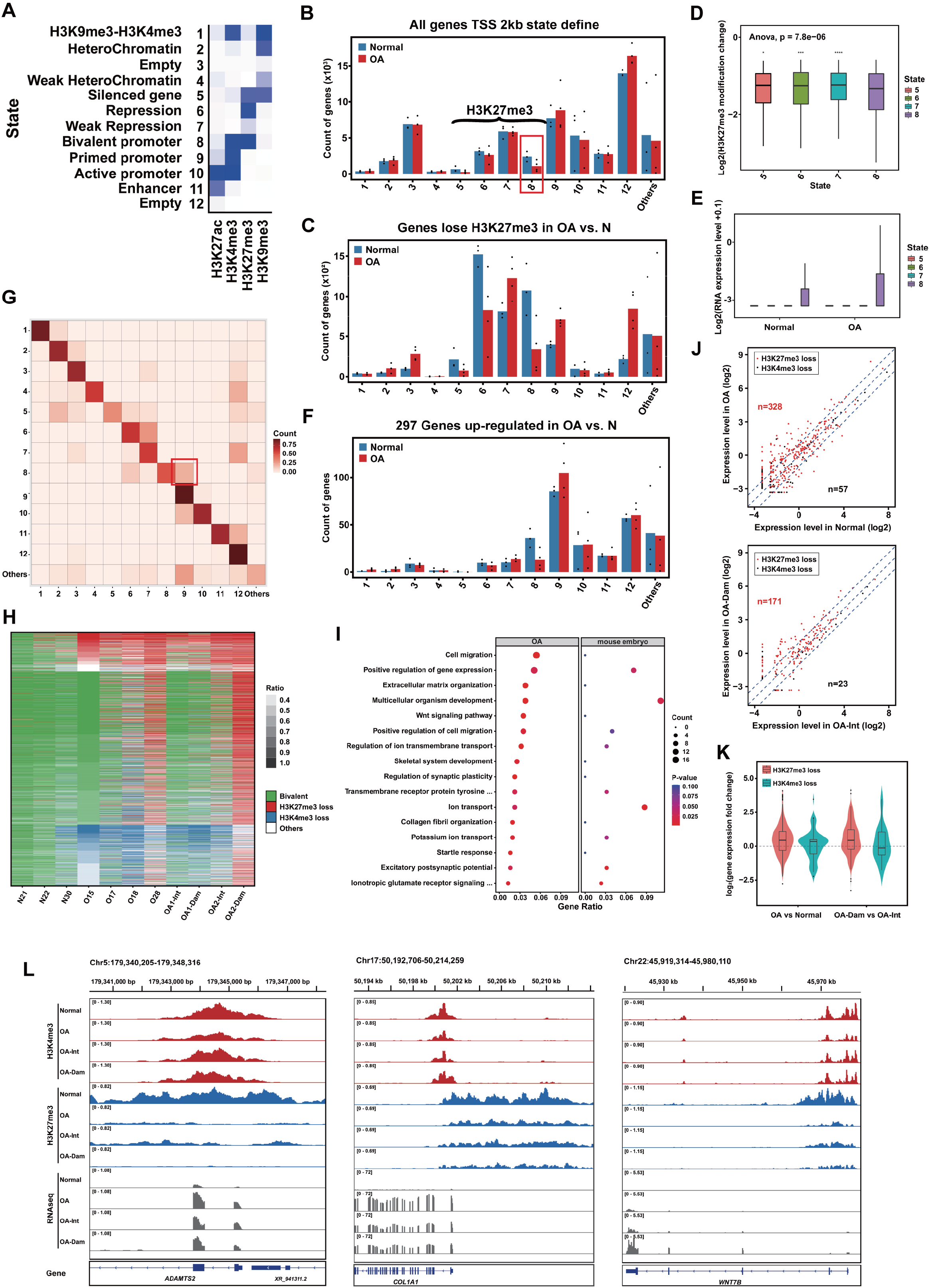
H3K27me3 loss in bivalent promoters up-regulate gene expression in OA. (A) Chromatin state definitions (emission probabilities) of a 12-state ChromHMM model based on the Normal, OA, OA-intact and OA-damaged data sets. (B) Promoter state define of all genes in each Normal and OA sample by the chromatin states of 2kb region around TSS. (C) Promoter state distribution of genes that lose H3K27me3 in OA vs. Normal. (D) Box plot showing the H3K27me3 changes of genes that lose H3K27me3 in OA vs. Normal with promoter state 5-8 in Normal. The significance of difference among all states was calculated by ANOVA test, and the significance of difference between state 8 and other states was determined by two side t test. (E) Box plot showing the RNA expression changes of genes that lose H3K27me3 in OA vs. Normal with promoter state 5-8 in Normal. (F) Promoter state distribution of up-regulated genes in OA vs. Normal. (G) Promoter state transitions of all genes from normal to OA. The transition ratio was normalized by the count of genes of each state in Normal. (H) Heatmap showing the K4/K27 bivalent promoters in normal samples and their state changes in OA, OA-Intact and OA-Damaged samples. (I) GO terms with the highest enrichment of bivalent genes which lose H3K27me3 in OA and their enrichment score of bivalent genes that lose H3K27me3 during endochondral ossification. (J) Dot plot showing the expression level of bivalent genes lose either H3K27me3 or H3K4me3 in OA vs Normal (up) and OA-Damaged vs OA-Intact (down). (K) Box/Violin plot showing the expression changes of bivalent genes lose either H3K27me3 or H3K4me3 in OA vs Normal (left) and OA-Damaged vs OA-Intact (right). (L) Examples of bivalent genes which lose H3K27me3 and gain expression during the progression of OA.

To find out if the loss of H2K27me3 has preference for bivalent promoters, we looked into the promoter states of about 5000 genes that lost H3K27me3 in OA versus Normal (Fig 1D). However, both bivalent (state 8) and repressive (state 6) genes exhibited significant enrichment in Normal (Fig 3C), with about a half of all genes in each state lost H3K27me3. Accordingly, genes in state 8 and 6 decreased significantly from Normal to OA (Fig 3C). Genes in state 9 and 7 increased in OA, which may result from the loss of H3K27me3 in state 8 and 6. Next we plotted the Log_2_ fold change of H3K27me3 in OA versus Normal for genes in states 5 to 8 (Fig 3D). Interestingly, genes in state 8 showed significantly more H3K27me3 down-regulation than other three groups. What’s more, the overall gene expression level of state 8 exhibited a significant up-regulation from Normal to OA, whereas genes in state 5-7 did not express in both Normal and OA (Fig 3E). These results suggested that H3K27me3 loss occurred mainly on genes of state 8 and 6. While only state 8, namely bivalent genes, resulted in the up- regulation of gene expression, which may due to the pre-exist of activating mark H3K4me3.

Next, we want to know the contribution of bivalent genes de-repression on the overall gene up-regulation. Among the 297 genes that up-regulated in OA versus Normal, genes with primed promoter (state 9) account for the largest proportion (Fig 3F). However, genes with active promoter (state 10) showed no increase in OA, we assumed that these genes used other transcriptional or post-transcriptional mechanism to increase expression level. Importantly, the most significant state change from Normal to OA is the loss of bivalent genes and increase of primed genes (Fig 3F).

State transition analysis verified the transition from bivalent genes in Normal to primed genes in OA (Fig 3G, state 8 to 9). Other obvious transitions include state 6 to 7, 7 to 12 and 5 to 2, which all attributed to the loss of H3K27me3. However, some bivalent genes also lost H3K4me3 to become repression (state 6).

### Bivalent genes derepression in OA resembles growth plate chondrocytes hypertrophy

We further analyzed states transitions of the 2346 bivalent genes in each sample (Fig 3H). A lot of bivalent genes lost H3K27me3 or H3K4me3 in OA. Especially, some OA samples exhibited an overall H3K27me3 loss (O28, O27-Dam). Paired samples also showed more H3K27me3 loss in Dam versus Int. Bivalent genes lost H3K27me3 in OA are enriched in GO terms that include cell migration, extracellular matrix organization, Wnt signaling pathway and ion transport (Fig 3I). Bivalent domains are deemed to poise developmental genes for future activation, whereas articular chondrocytes are meant to persist to form permanent cartilage. Originating from the same cartilage mass, growth plate chondrocytes undergo proliferation, hypertrophy and apoptosis, which promotes degradation and calcification of cartilage matrix to drive bone growth. OA pathogenesis commonly includes a switch from the articular to the growth plate chondrocyte phenotype, and re-initiation of proliferation and hypertrophy (Pitsillides & Beier, 2011).

We wondered whether the bivalent genes need to be activated during the proliferation and hypertrophy of growth plate chondrocytes but remain as a developmental legacy feature in articular chondrocytes. Re-analysis of the epigenetic data from a mouse study (Wuelling *et al*, 2021) revealed 162 bivalent genes in proliferating chondrocytes that lost H3K27me3 in hypertrophic chondrocytes (Fig EV4, C to E) (Table S3). GO analysis revealed enrichment in several terms like multicellular organism development and ion transport (Fig 3I). Overlapping GO terms between OA and growth plate chondrocytes hypertrophy suggested the similarity between the two process.

Next, we defined the bivalent genes that lose H3K27me3 or H3K4me3 in OA versus Normal and Damaged versus Intact. Apparently, the vast majority of the altered bivalent genes lose H3K27me3 and only a very small number lose H3K4me3 (Fig 3J). Furthermore, the bivalent genes lost H3K27me3 tend to obtain higher expression level in both OA versus Normal and Damaged versus Intact (Fig 3, J and K). Typical bivalent genes that lose H3K27me3 and gain expression in OA were shown (Fig 3L). These data suggest that H3K27me3 loss in bivalent promoters up-regulate gene expression in OA articular chondrocytes, which is consistent to the hypertrophy and ossification of growth plate chondrocytes.

### Inflammation induced H3K27 demethylase expression in OA chondrocytes

Next, we wondered what caused the loss of H3K27me3 on bivalent promoters in OA. H3K27me3 are added by the PRC2 complex that consist EZH1/2, EED and SUZ12, and removed by KDM6A and KDM6B (Xiang *et al*, 2007; Hong *et al*, 2007; Agger *et al*, 2007; De Santa *et al*, 2007). However, the RNA expression of these methyltransferases and demethylases in our RNA-seq dataset showed no significant difference in OA versus Normal and Damaged versus Intact (Fig EV5). Inflammation plays an important role in the development of OA. Proinflammatory cytokines like IL-1β downregulate the synthesis of major extracellular matrix (ECM) components and induce the production of proteolytic enzymes of chondrocytes. Thus, IL-1β is widely used to create a cellular model of OA. Interestingly, IL-1β treatment increased the expression of H3K27me3 demethylases in human chondrocytes from OA patients (Fig 4A), with a higher induction ratio of KDM6B than KDM6A. It has been reported that KDM6B (JMJD3) contributes to the control of gene expression in LPS-activated macrophages (De Santa *et al*, 2007, 2009) and is involved in inflammatory diseases. More importantly, KDM6B has been reported to promote chondrocyte proliferation and hypertrophy in endochondral bone formation (Zhang *et al*, 2015, 3). These results suggested KDM6B as a candidate for H3K27me3 loss in OA. However, KDM6B showed no statistic difference in human normal and OA cartilage by immunofluorescence (IF) staining (Fig 4, B and C), in which KDM6B positive cells were around 50% in both groups. However, acute inflammation is contraindicated for knee replacement. So, the OA cartilage samples we collected here were inflammation quiescent and may not exhibit KDM6B up-regulation. A time course that simulating the transient inflammation in OA patients detected the fall back of KDM6B after IL-1β removal (Fig 4D). We then established a rat surgical instability OA model to examine KDM6B in cartilage and synovial inflammation by *H&E* staining. OA synovium exhibited typical hyperplasia and plenty of inflammatory cells in both 4w and 8w (Fig 4E). Interestingly, KDM6B positive cells raised from sporadic in shame to about 50% in OA (Fig 4F). These data suggest that inflammation induces the expression of KDM6B, which may be responsible for the H3K27me3 loss in OA chondrocytes.

**Figure 4.**
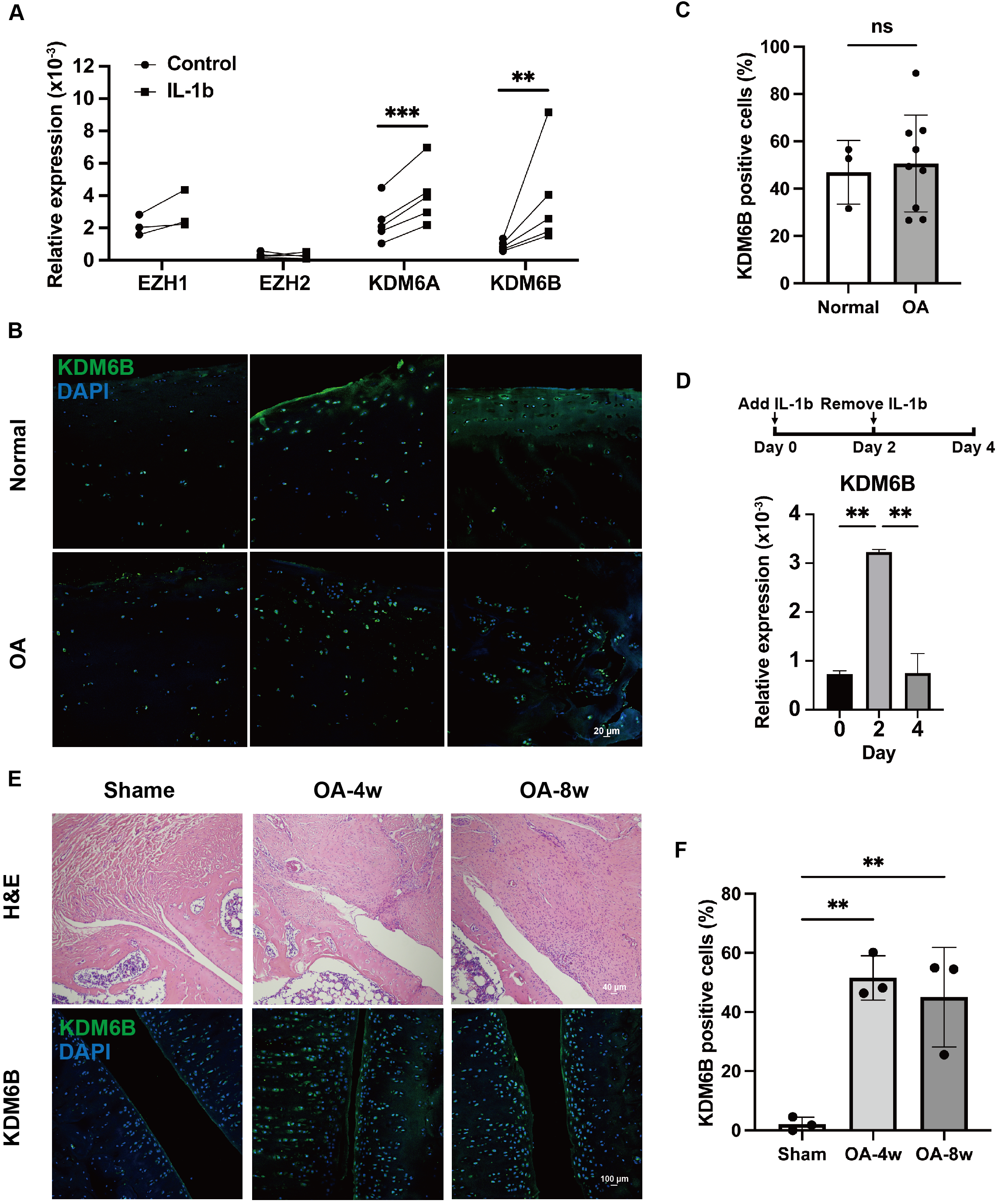
Inflammation induced H3K27 demethylase expression in OA chondrocytes. (A) The RNA expression level of H3K27me3 methylases and demethylases in paired control and IL-1β treated human chondrocytes from OA patients (n=3 for EZH1, n=5 for others). Data were analyzed by two-tailed ratio paired t test. (B) Immunofluorescence for KDM6B in human normal and OA cartilage. (C) Quantification of KDM6B-positive cells in human normal (n=3) and OA (n=9) samples. Data were analyzed by two-tailed unpaired t test and are shown as means ± SD. (D) The RNA expression level of KDM6B during a time course of IL-1β addition and removal. The same sample was measured repeatedly (n=3). Data were analyzed by ordinary one-way ANOVA followed by Tukey’s multiple comparisons test and are shown as means ± SD. (E) H&E staining for synovium and immunofluorescence for KDM6B in rat ACLT- DMM OA models. (F) Quantification of KDM6B-positive cells in rat control (n=3) and OA (n=3) samples. Data were analyzed by ordinary one-way ANOVA followed by Tukey’s multiple comparisons test and are shown as means ± SD. **P < 0.01,***P < 0.001.

### KDM6 inhibitor rescued inflammation-induced H3K27me3 loss and abnormal gene expression

We tested whether the inflammation induction of KDM6B causes H3K27me3 loss in human OA chondrocytes by CUT&Tag. It was found that IL-1β treatment resulted in a significant H3K27me3 decrease across gene bodies on the whole genome (Fig 5A). Interestingly, 48hr after the withdraw of IL-1β, with the expression level of KDM6B fell back (Fig 4D), the H3K27me3 level didn’t recover. This may explain the contradiction of H3K27me3 loss and normal KDM6B level in OA patients (Fig 4, B and C). Acute inflammation in OA patients induce the transient expression of KDM6B, which may result in H3K27me3 loss that could persist even after the acute inflammation.

**Figure 5.**
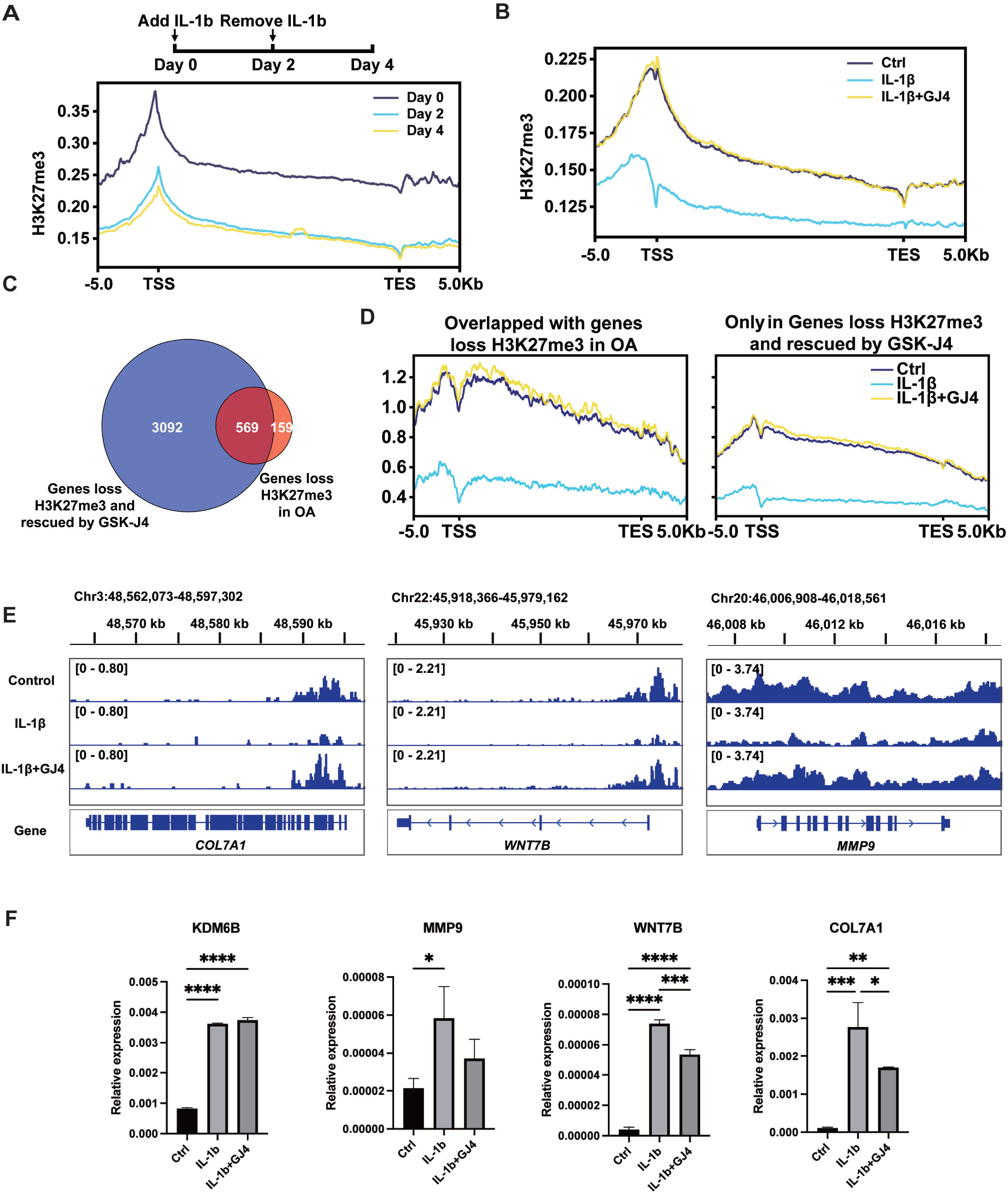
Demethylase inhibitor rescued inflammation induced H3K27me3 loss and abnormal gene expression. (A) Tag density of H3K27me3 across the gene bodies (+/-5kb) genome-wide in human OA chondrocytes during a time course of IL-1β addition and removal. (B) Tag density of H3K27me3 across the gene bodies (+/-5kb) genome-wide in human OA chondrocytes treated with IL-1β and GSK-J4 (GJ4). (C) Venn diagram of genes which lose H3K27me3 upon IL-1β treatment and restored H3K27me3 by GSK-J4 and genes loss H3K27me3 in OA samples. (D) H3K27me3 tag density of the overlapped 569 genes (left) and the other 3092 genes (right) in Fig 3C. (E) Examples of H3K27me3 distributions around genes which lose H3K27me3 upon IL-1β treatment and restored H3K27me3 by GSK-J4. (F) Relative expression of genes which lose H3K27me3 upon IL-1β treatment and restored H3K27me3 by GSK-J4. The same sample was measured repeatedly (n=3). Data were analyzed by ordinary one-way ANOVA and are shown as means ± SD. *P < 0.05; **P < 0.01; ***P < 0.001; ****P < 0.0001.

To verify the importance of KDM6B in this process, we used a selective KDM6 inhibitor, GSK-J4 (Kruidenier *et al*, 2012). Surprisingly, combined utilization of GSK-J4 and IL-1β completely inhibited this H3K27me3 loss (Fig 5B). 3661 genes were identified to lose H3K27me3 upon IL-1β and simultaneously rescued by GSK-J4, which contain 569 genes of the previously found 728 genes that lose H3K27me3 in OA versus Normal and Damaged versus Intact (Fig 5C). Furthermore, the overlapped 569 genes showed higher H3K27me3 in control and GSK-J4 rescued samples than the rest 3092 genes (Fig 5D), suggesting the consistence between *in vivo* and *in vitro* conditions. Typical genes that lost H3K27me3 upon IL-1β and restored H3K27me3 by GSK-J4 were shown (Fig 5E). Accordingly, their gene expression level increased upon IL-1β and were repressed by GSK-J4 (Fig 5F). Therefore, a key role of KDM6B was proposed in inflammation induced H3K27me3 loss and abnormal gene expression in OA chondrocytes (Fig 6).

**Figure 6.**
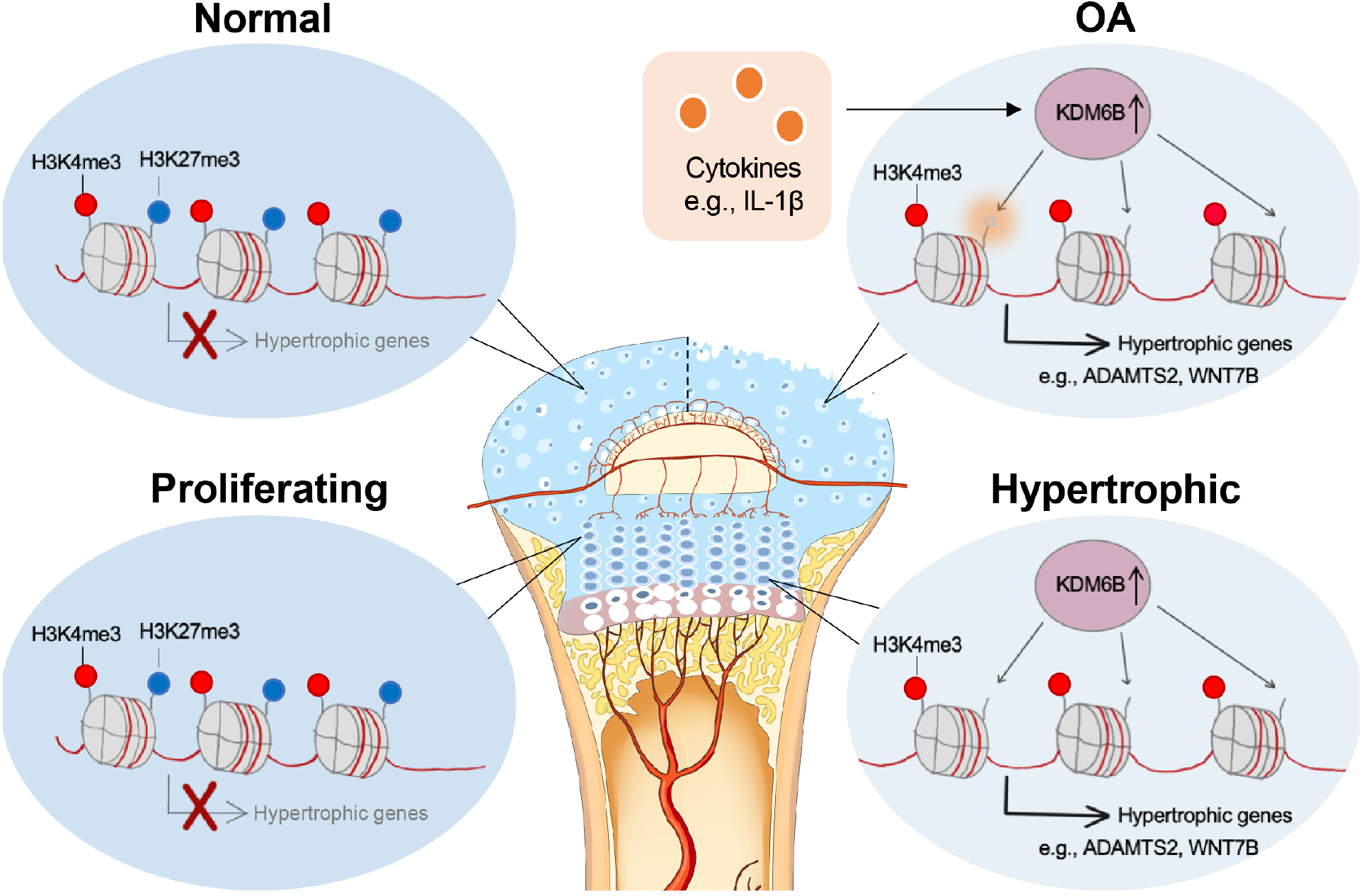
Working model: Inflammation induced epigenetic activation of bivalent genes in osteoarthritic cartilage. During the osteogenesis of growth plate cartilage, hypertrophy related genes were occupied by both H3K27me3 and H3K4me3 due to the need for rapid activation. However, in its homologous articular chondrocytes, hypertrophic genes remained in a bivalent state. In normal conditions, these bivalent genes maintain transcriptional inhibition; In OA, inflammatory cytokines such as IL-1β induce the expression of H3K27me3 demethylase KDM6B to remove H3K27me3 thus facilize the expression of hypertrophic genes, which promote the degradation of cartilage matrix and leads to OA.

Altogether, our data revealed the loss of H3K27me3 and de-repression of bivalent genes that caused chondrocyte hypertrophy in OA. Inflammation induced the expression of the H3K27me3 demethylase KDM6B, thus provides epigenetic mechanism for IL-1β induced cartilage destruction.

## Discussion

Our study, for the first time, provides a genome-wide histone modification map for OA chondrocytes. The most significant change in OA is the loss of repressive mark H3K27me3 on promotors, which in turn lead to up-regulation of OA related genes. Surprisingly, in normal chondrocytes plenty of genes are bivalent, allowing easily activation upon H3K27me3 loss in OA, which resemble hypertrophy related genes in growth plate chondrocytes. This finding provides novel epigenetic evidence for the developmental explanation for OA. Further investigation revealed that H3K27me3 demethylase KDM6B is highly expressed in a rat OA model and also induced by IL-1β in human OA chondrocytes. IL-1β treatment leads to significant H3K27me3 loss and up- regulation of OA related genes. While a KDM6B inhibitor, GSK-J4, fully restores H3K27me3 and partially down-regulates gene expression, which serves as an attractive therapeutic strategy for OA.

Several studies have addressed the importance of H3K27me3 methyltransferases and demethylases in cartilage development and OA. Loss of PRC2 components Ezh2 or Eed in murine chondrocytes accelerate hypertrophic differentiation and stimulates an osteogenic transcriptional program (Camilleri *et al*, 2018, 2; Mirzamohammadi *et al*, 2016), highlighting the importance of H3K27me3 in suppressing Wnt and TGF-β signaling. Another study found that EZH2 was highly expressed in a subpopulation of OA patients and ameliorates OA by inhibiting hypertrophy (Du *et al*, 2020, 2), consistent with our findings. Before the finding in endochondral bone formation (Zhang *et al*, 2015, 3), KDM6B is known to be an inducible enzyme that plays key roles in development (Agger *et al*, 2007; Estarás *et al*, 2012) and inflammatory response (De Santa *et al*, 2007, 2009). The effect of KDM6B in OA is controversy. One study reported that knockdown of Kdm6b in mouse chondrocytes leads to accelerated OA progression, with the number of Kdm6b-positive chondrocytes lower in both mice and human OA cartilage samples (Dai *et al*, 2017). While other studies reported an increased expression of KDM6B in OA chondrocytes, and GSK-J4 prevented the expression of pro-inflammatory cytokines and catabolic enzymes (Yapp *et al*, 2016, 3; Jun *et al*, 2020). Furthermore, the latter study demonstrated a cartilage protection effect of GSK- J4 in a surgically induced mouse OA model (Jun *et al*, 2020).

Our research on human OA cartilage samples showed a significant loss of H3K27me3 genome-wide and especially on hypertrophic genes, supporting an OA promoting effect for KDM6B. We also found KDM6B up-regulation in a rat OA model and IL-1β treated human OA chondrocytes, but not in human OA specimen. In addition to the inflammation quiescent explanation we discussed above, another possibility is the dysregulation of cofactors that bring KDM6B to specific genome locus. As GSK-J4 is an inhibitor of both KDM6B and KDM6A, mapping global targets of this two demethylases and putative cofactors in OA cartilage can provide us better insights into the putative candidature of GSK-J4 and related molecules for OA therapeutics. In addition, we found an increase of H3K4me3 in intergenic regions indicating the loss of preciseness in epigenetic regulation that requires further investigation.

In this study, we found that inflammation induced KDM6B lead to a significant decrease of H3K27me3 which lead to the activation of bivalent genes in OA. Our results suggest an inherited epigenetic signature that makes articular chondrocytes prone to hypertrophy and ossification and contribute to a promising epigenetic therapy for OA.

## Materials and Methods

### Human articular cartilage samples

Experiments involving human subjects were performed according to the Ethics Committee on Biomedical Research, West China Hospital of Sichuan University. Informed consent was obtained from patients. OA cartilage tissues were obtained from patients undergoing total knee arthroplasty due to end-stage OA at West China Hospital. Normal cartilage tissues were obtained from individuals undergoing amputation (without OA). The whole layer of cartilage from medial and lateral femoral condyle were cut into small pieces and digested by 4mg/ml PRONASE Protease (Millipore, 53702) for 1hr followed by 1mg/ml collagenase (Nordmark, S1746502) in DMEM/F12 with 10% FBS overnight. The dissociated chondrocytes were then used for CUT&Tag and RNA-seq library construction.

### CUT&Tag library construction and sequencing

CUT&Tag libraries were constructed as previously described (Kaya-Okur *et al*, 2020). In brief, chondrocytes were lysed to acquire nuclei and bind to Concanavalin-coated magnetic beads (Polysciences, 86057). Beads were incubated successively with the primary antibody (Anti-H3K4me3, Abcam, ab8580; Anti-H3K27me3, Active motif, 39055; Anti-H3K27ac, Abcam, ab177178; Anti-H3K9me3, Abcam, ab176916), the secondary antibody (Goat Anti-Rabbit IgG(H+L), Abclonal, AS070) and pG-Tn5 (Vazyme, S602) loaded with mosaic-end adapters. Tn5 was activated by addition of Mg^2+^. DNA was released in a small volume of SDS. Samples were enriched by PCR amplification (NEBNext, 2X PCR premix, M0541) followed by magnetic bead cleanup (Vazyme, N411) and sent for paired-end 150-bp sequencing performed on an Illumina HiSeq 2500 platform.

### Rat OA model

Experiments involving animal subjects were performed in accordance with Institutional Animal Care and Use Committee (IACUC) of the Sichuan University-approved protocols. Six-week-old male SD rats were purchased from Chengdu Enswille Biotechnology Co., LTD. Two weeks after adaptation, the OA model was established by anterior cruciate ligament transection (ACLT) plus destabilization of the medial meniscus (DMM) under isoflurane anesthesia. Briefly, the skin was cut approximately 2cm along the medial side of the knee joint, and the tissue was subsequently separated until the patellar ligament was exposed. The joint capsule was opened medial to the patellar ligament. After cutting the anterior cruciate ligament, the anterior horn of the medial meniscus was carefully removed along the medial edge of the tibial plateau (sham operation group only opened the joint capsule without other surgery), and finally the joint capsule, muscle and skin were sutured. At four or eighth week after surgery, rats were sacrificed under isoflurane anaesthetization.

### Cell culture and treatment

The dissociated chondrocytes were cultured in DMEM/F12 with 2% FBS. For time course that simulating the acute inflammation, 5ng/ml IL-1β was added for 2days and then removed for another 2 days. For the rescue experiment, 5ng/ml IL-1β alone or in combination with 5uM GSK-J4 (abmole, M5149) were added for 48hrs.

### CUT&Tag data processing

Read quality of the raw sequences was verified with fastqc (0.11.9). All sequence reads were then aligned to the hg38 build of the human reference genome by bowtie2 (2.3.5.1) using --end-to-end --very-sensitive --no-mixed --no-discordant --phred33 -I 10 -X 700 parameters. Duplication rate of each sample was checked with Picard tools (2.3.1). Mapped reads were normalized to 1 million to generate histone modification enrichment around genes with deeptools (3.5.1) (Ramírez *et al*, 2016). For H3K4me3, H3K27me3, H3K27ac and H3K9me3, to assess data reproducibility between different samples, correlation analysis of normalized mapped reads counts in each 500-bp bins across the whole genome was calculated.

All CUT&Tag peaks were called by SEACR (1.3) (Meers *et al*, 2019) with parameters 0.01 non stringent. Subsequently, these identified peaks were classified into five categories with the following order: promoter, gene body, enhancer, region 10kb nearby TSS and intergenic region. To calculate the differential peaks of particular histone modification, we combined all the mapped reads of each sample to identify “common” peaks of this histone modification. The tags abundance of each peak in Normal, OA, OA-Int and OA-Dam sample was quantified independently and limma was used for differential expression analysis. The peaks exhibiting tags abundance foldchange >2 or <0.5 and P-value <0.05 over OA to Normal or OA-Dam to OA-Int were defined as up- regulated or down-regulated peaks. To calculate the differential genes of particular histone modification, we calculated the tags abundance on all gene promoters, which were defined as +- 2kb region around the TSS of each gene, in each sample. The genes exhibiting tags abundance foldchange >2 or <0.5 and P-value <0.05 over OA to Normal or OA-Dam to OA-Int were defined as up-regulated or down-regulated genes. The mapped CUT&Tag reads and identified peaks were visualized on the Integrative Genomics Viewer (IGV).

### RNA-seq data processing

Hisat2 (2.1.0) software was used to align the clean sequencing reads to the human reference genome (hg38) with parameters (--no-unal --rna-strandness FR --fr). Expression levels for all genes were quantified to fragments per kilobase million (FPKM) using Stringite (2.2.0) with default parameters. Subsequently, edge R was used for differential expression analysis. The genes with expression level fold change >2 or <0.5 and P-vaule <0.05 was defined as up-regulated genes or down-regulated genes respectively. Gene Ontology analysis were performed using DAVID (Sherman *et al*, 2022).

### Histone modifications and its correlation with gene expression

To quantify correlation of histone modification levels with gene expression value, tags in identified peaks which overlapped with gene promoters were summed and normalized for each gene to represent the modification levels. The genes were then ranked and binned by its modification levels. Each bin of genes (100) is represented by a dot on the plot, with the arithmetic mean of histone modification and gene expression levels to stand for the histone modification and gene expression level of the bin.

To quantify correlation of histone modification changes with gene expression changes, the modification levels of each gene was also calculated as the same manner. The change of modification level and gene expression level was calculated as the logarithm of the ratio of the normalized modification levels and gene expression levels in OA versus Normal or OA-Dam versus OA-Int. The genes were then ranked and binned by its modification changes, with the average of their expression changes and modification changes to represent one dot in the plot.

### Defining epigenetic states with ChromHMM

The mapped CUT&Tag reads were used as input for ChromHMM (1.24) (Ernst & Kellis, 2012). The whole genome was segmented into non-overlapping bins of 200 bp. A multivariate Hidden Markov Model was used to model the combinatorial and spatial patterns from H3K4me3, H3K27me3, H3K27ac and H3K9me3 markers in each bin. Based on their estimated log-likelihoods, the 12-state ChromHMM model was selected. According to the combinatorial patterns of 4 histone markers of each state, we defined these states as H3K9me3-H3K4me3, HeteroChromatin, Empty, Weak HeteroChromatin, Silenced gene, Repression, Weak Repression, Bivalent promoter, Primed promoter, Active promoter, Enhancer and empty. To assess the robustness of trained model between different samples, we used kappa analysis to evaluate the state agreement between Normal, OA, OA-Int and OA-Dam subsets. The fold enrichment of each ChromHMM state was also quantified at Refseq gene, Refseq exon, RefSeq TSS, RefSeq TSS, +-2kb region around RefSeq TSS and CpG islands.

### Analysis of ChromHMM bivalent promoter state transition

For ChromHMM bivalent promoter state transition analysis of Normal to OA and OA- Int to OA-Dam, we calculated the bivalent promoter state occurrence in +- 1kb region around TSS (10 ChromHMM bins) of each gene. The genes with at least 7 bivalent promoter state in at least 2 of 3 Normal samples was defined as bivalent genes. We then count and plot heatmap to show the occurrence of ChromHMM state transition in +- 1kb region around TSS of each bivalent gene in OA, OA-Int and OA-Dam subsets. For each bivalent gene, “H3K27me3 loss” was happened in the subset if there was at least 4 ChromHMM primed promoter or active promoter state occurred, “H3K4me3 loss” was happened in the subset if there was at least 4 ChromHMM repression state occurred, “bivalent” was happened in the subset if there was still at least 6 ChromHMM bivalent promoter state occurred.

To figure out the genes with H3K27me3 loss and H3K4me3 loss in the process of Normal to OA or OA-Int to OA-Dam, we first defined the states transition of each bin according to the occurrence in all samples. The bins with at least 2 ChromHMM primed promoter or active promoter state in 4 OA subsets were defined as H3K27me3 loss bins in OA, the bins with at least 1 ChromHMM primed promoter or active promoter state in 2 OA-Int or OA-Dam subsets were defined as H3K27me3 loss bins in OA-Int or OA- Dam, the bins with at least 2 ChromHMM repression state in 4 OA subsets as H3K4me3 loss bins in OA, the bins with at least 1 ChromHMM repression state in 2 OA-Int or OA- Dam subsets as H3K4me3 loss bins in OA-Int or OA-Dam. The genes with at least 40% bivalent bins occurred H3K27me3 loss or H3K4me3 loss was defined as H3K27me3 loss genes or H3K4me3 loss genes. The expression levels in Normal, OA, OA-Int and OA-Dam and expression changes between them were then counted and plotted.

### Statistical analysis

Analyses were performed using GraphPad Prism 9.0 (GraphPad Software, USA). All data were presented as means ± SDs. Statistical comparisons of two groups were performed by two-tailed ratio paired or unpaired t test. Multiple comparisons were made using ordinary one-way ANOVA followed by Tukey’s multiple comparisons test. Statistical significance (*P < 0.05; **P < 0.01; ***P < 0.001; ****P < 0.0001) was indicated in figures.

## Data Availability

The CUT&Tag and RNA-seq data generated in this study have been deposited in the NCBI's Sequence Read Archive (SRA) database under accession code PRJNA930583. The mouse PC and HC ChIP-seq data was downloaded through the GEO accession numbers. All code required for data analyses and figure preparation are available at https://github.com/luciferase1234/Inflammation-induced-epigenetic-activation-of-bivalent-genes-in-osteoarthritic-cartilage. All data are available in the main text or the supplementary materials.

## Acknowledgments

We are grateful to the IT Center of west china hospital for supplying the computing resources. We thank Professor Hongjie Shen for his advice and criticism. This work was supported by National Natural Science Foundation of China (81874027, U22A20280); Science & Technology Department of Sichuan Province (2023NSFSC1607, 2021YFSY0003,2022YFS0051,2022NSFSC1312); Shenzhen-HongKong Institute of Brain Science-Shenzhen Fundamental Research Institutions (312200102); Clinical Research Incubation project of West China Hospital of Sichuan University (2021HXFH036).

## Author contributions

HD conceived the project, designed and performed the experiments, analyzed the data, and wrote the manuscript. YZhang co-conceived the project, did all the bioinformatics analysis and prepared figures. XYu and DW performed cell isolations. XYou performed immunofluorescence microscopy and helped with Cut&Tag. ZLuo and YC performed animal experiments. HL and ZLiao participated in clinical sample acquisition. B-SD, YZhao and YW provided suggestions and revised the manuscript. KX drawed part of the working model. FY, FG, NN and JZ revised the manuscript. ZZ and SH supervised the work and revised the manuscript.

## Expanded View Figure legends

**Figure EV1.**
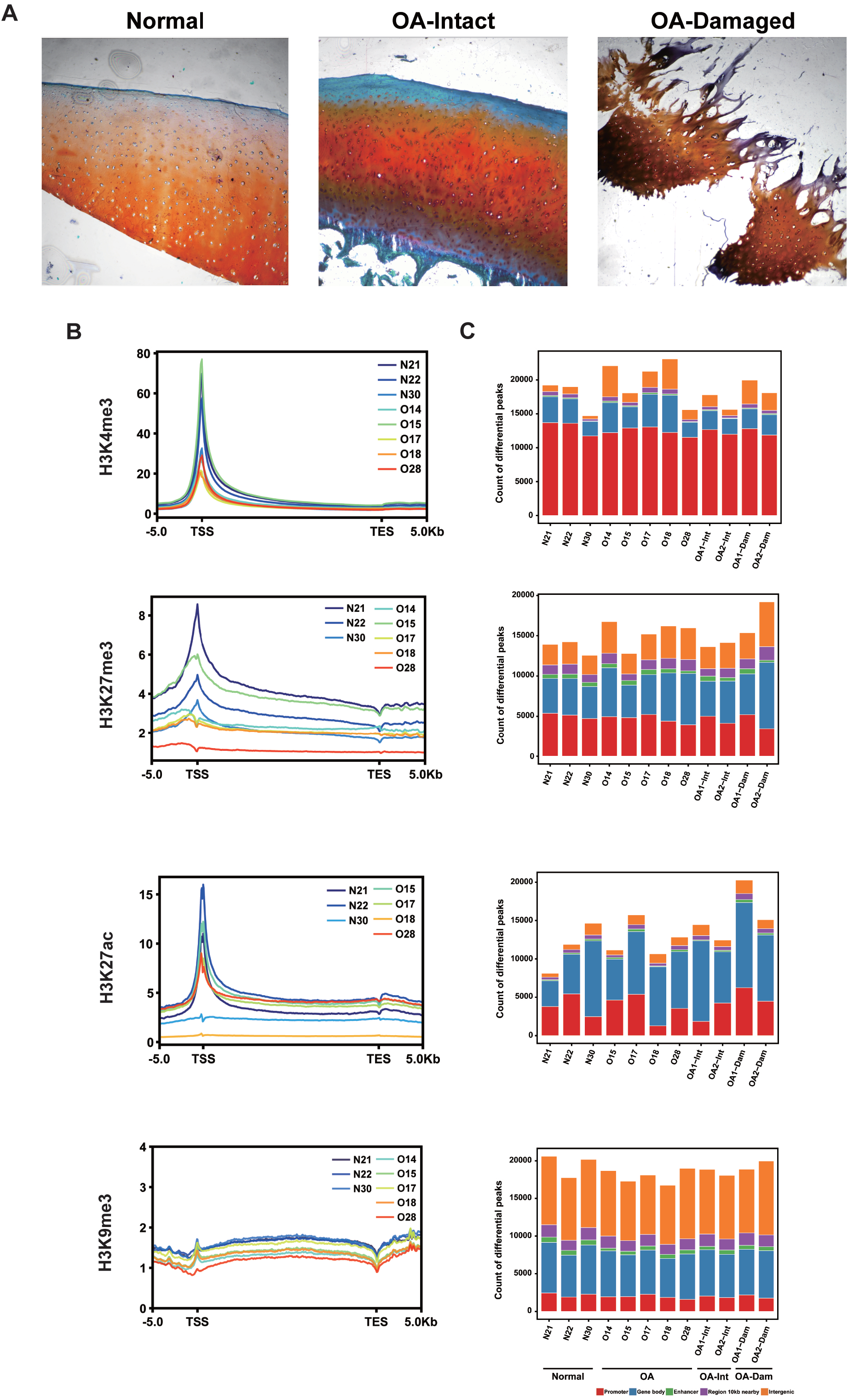
Histone modification profiling of each normal and OA sample. (A) Safranin O-Fast Green staining of human femoral condylar cartilage used for histone modification profiling. (B) Tag density of each histone mark across the gene bodies (+/-5kb) in each Normal and OA samples. (C) Genome-wide peak distribution of each modification in each sample. The numbers of peaks in promoter, gene body, enhancer, 10kb nearby gene body, and intergenic regions are shown. (D) Correlation map of each sample for the four histone marks.

**Figure EV2.**
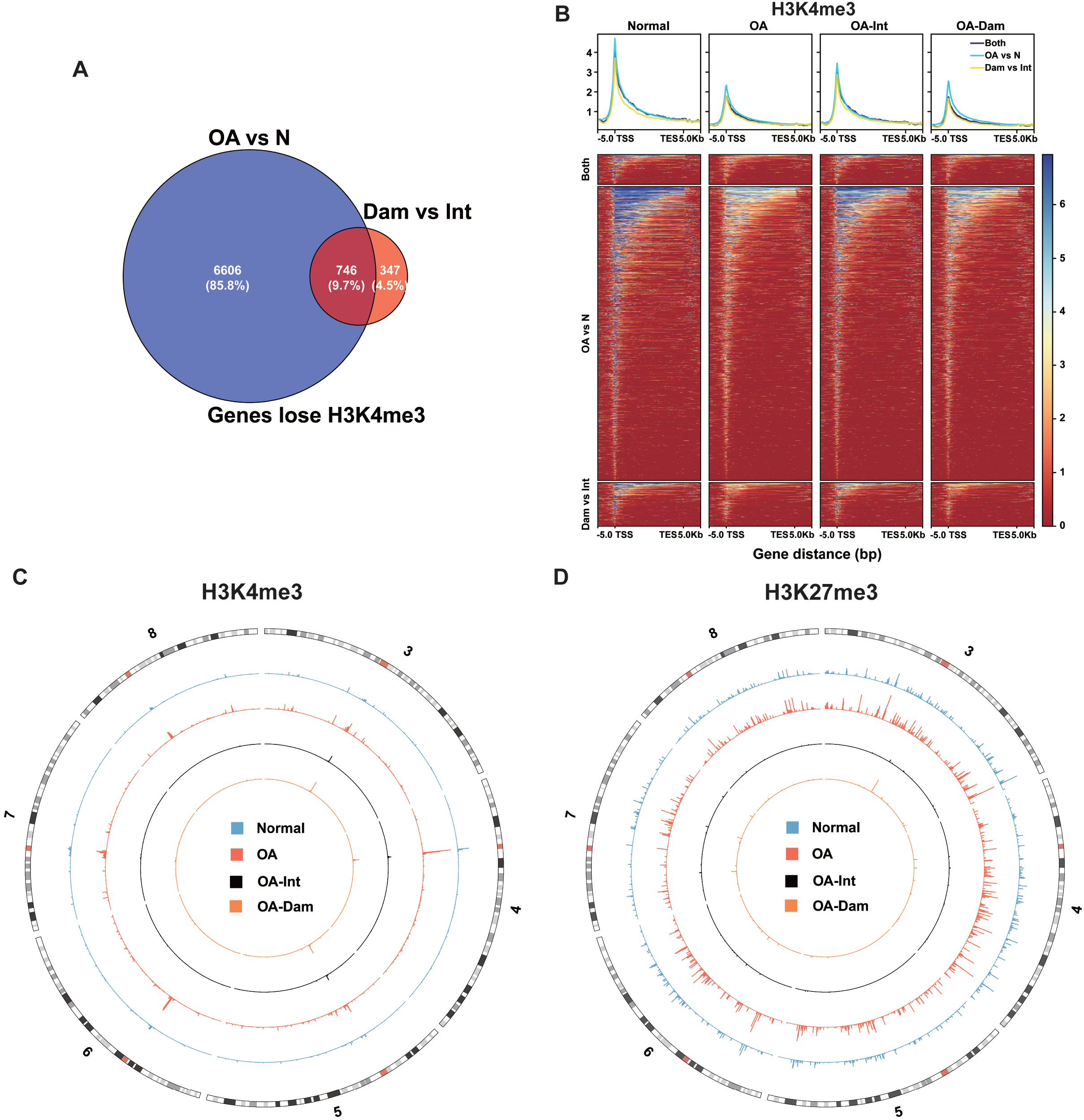
Differences of H3K4me3 and H3K27me3 in normal and OA. (A) Venn diagram of genes which lose H3K4me3 in OA vs. Normal and OA-Damaged vs. OA- Intact. (B) Tag density and heatmap of H3K4me3 across the gene bodies (+/-5kb) of genes which loss H3K4me3 in OA vs. Normal, OA-Damaged vs. OA-Intact and both. (C) Chromosome distribution of up-regulated H3K4me3 peaks outside genes in OA vs. Normal and Dam vs. Int respectively. (D) Chromosome distribution of up-regulated H3K27me3 peaks outside genes in OA vs. Normal and Dam vs. Int respectively.

**Figure EV3.**
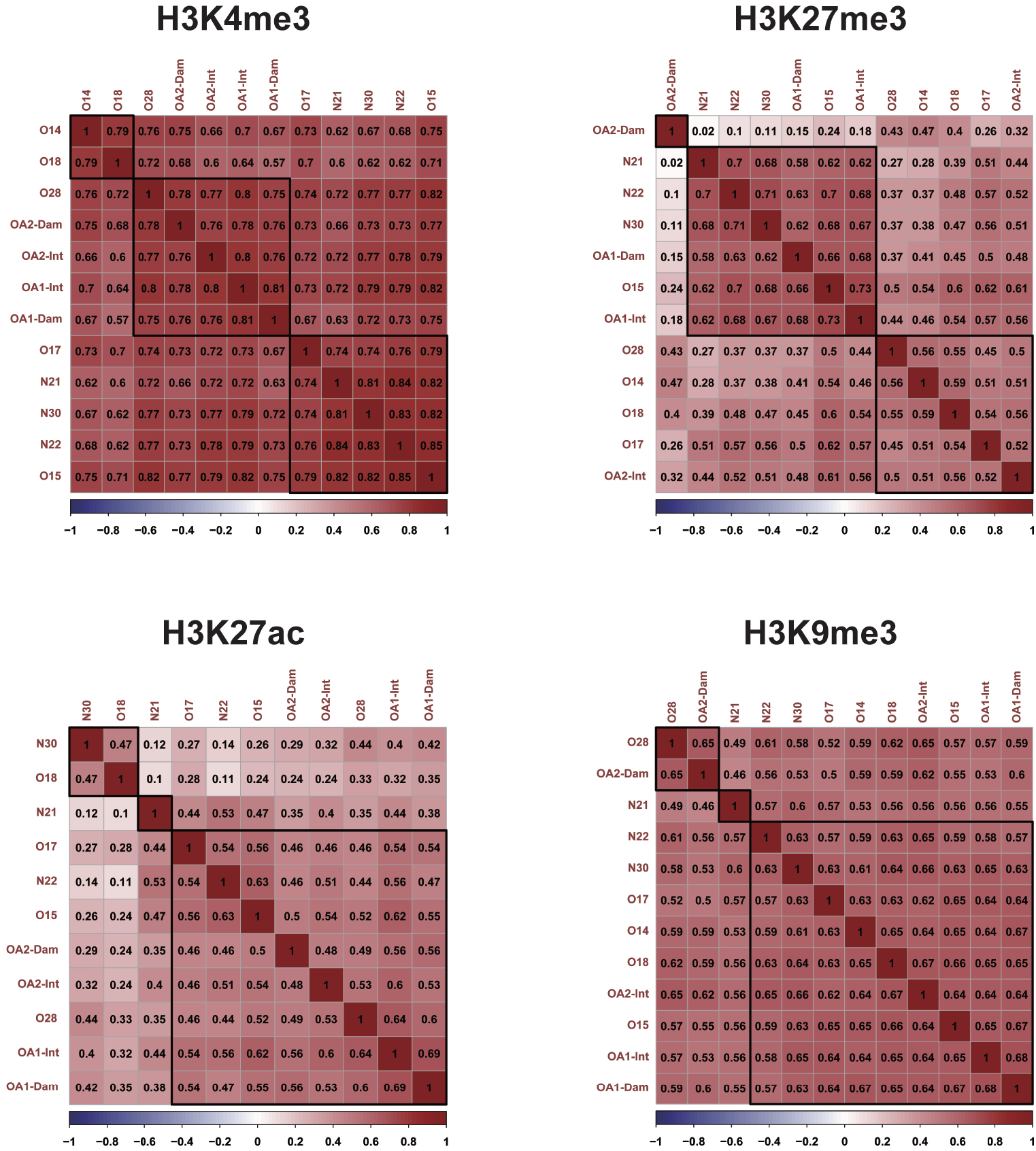
Correlation map of each sample for the four histone marks.

**Figure EV4.**
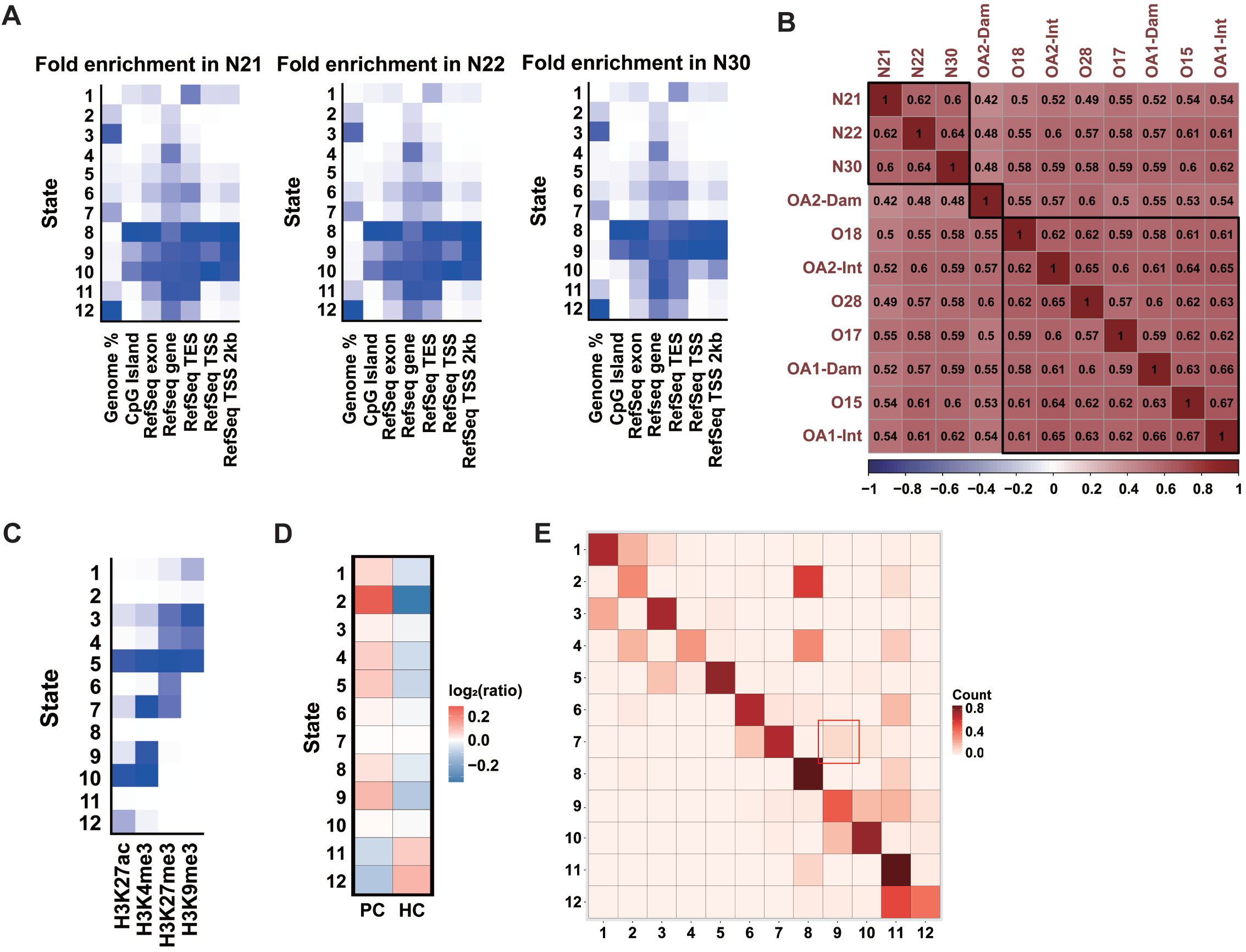
ChromHMM analysis of human and mice datasets. (A) Enrichment scores for chromatin states in genomic features for 3 normal samples. The first column shows the genome-wide percentage of occupancy for each state. Subsequent columns show enrichments for CpG islands, Refseq annotated exons, genes, transcription ends sites (TES), transcription start sites (TSS) and TSS + −2 kb regions. Each column is colored from 0 (white) to its maximum value (blue). (B) Heatmap showing the state agreement between Normal, OA, OA-Int and OA-Dam subsets. (C) Chromatin state definitions (emission probabilities) of a 12-state ChromHMM model based on the mouse PC/HC data sets. (D) The count of genomic bins in each state in mouse PC/HC data sets. (E) State transitions from PC to HC.

**Figure EV5.**
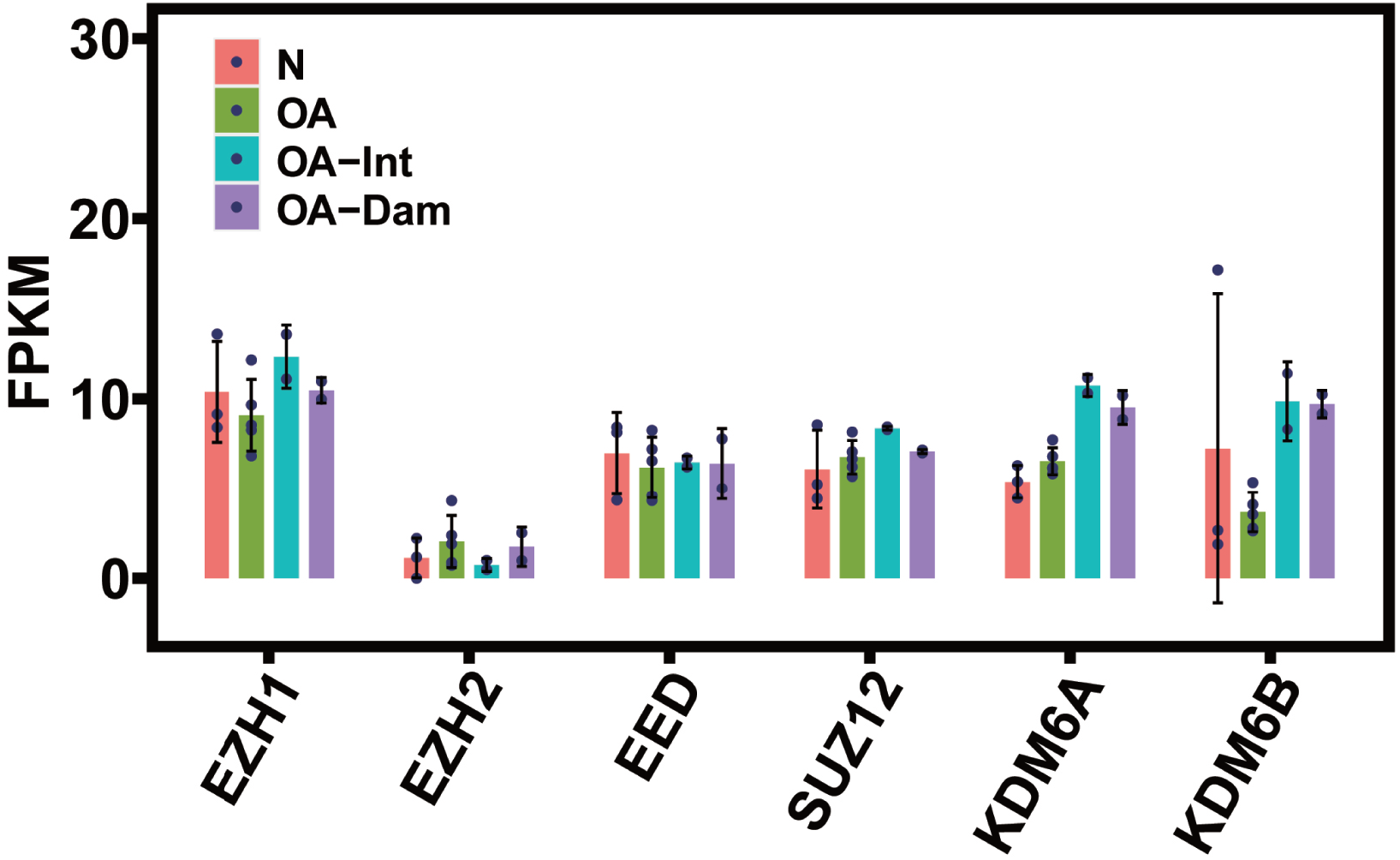
The expression of H3K27 methylation regulators in RNA-seq data from 4 groups.

**Table EV1. Quality control of CUT&Tag libraries**.

**Table EV2. Quality control of RNA-seq**.

**Table EV3. Bivalent genes in mouse proliferating chondrocytes that lost H3K27me3 in hypertrophic chondrocytes**.

